# Increasing activity after stroke: a randomized controlled trial of high-intensity walking and step activity intervention

**DOI:** 10.1101/2023.03.11.23287111

**Authors:** Elizabeth D. Thompson, Ryan T. Pohlig, Kiersten M. McCartney, T. George Hornby, Scott E. Kasner, Jonathan Raser-Schramm, Allison E. Miller, Christopher E. Henderson, Henry Wright, Tamara Wright, Darcy S. Reisman

**Affiliations:** University of Delaware, Department of Physical Therapy, Newark, DE, USA; University of Delaware, Biostatistics Core, Newark, DE, USA; University of Delaware, Biomechanics and Movement Science (BIOMS) program, Newark, DE, USA; Indiana University, Department of Physical Medicine and Rehabilitation, Indianapolis, IN, USA; University of Pennsylvania, Perelman School of Medicine, Philadelphia, PA, USA; Christiana Care Health System, Newark, DE, USA; Washington University School of Medicine, Program in Physical Therapy, St. Louis, MO, USA

**Keywords:** walking, stroke, high-intensity gait, step activity monitoring, treadmill

## Abstract

**Background:** Physical inactivity in people with chronic stroke profoundly affects daily function and increases recurrent stroke risk and mortality, making physical activity improvements an important target of intervention. We compared the effects of a high-intensity walking intervention (FAST), a step activity monitoring behavioral intervention (SAM), or a combined intervention (FAST+SAM) on physical activity (i.e., steps per day). We hypothesized the combined intervention would yield the greatest increase in steps per day.

**Methods:** This assessor-blinded multi-site randomized controlled trial was conducted at four university/hospital-based laboratories. Participants were 21-85 years old, walking without physical assistance following a single, unilateral non-cerebellar stroke of ≥6 months duration, and randomly assigned to FAST, SAM, or FAST+SAM for 12 weeks (2-3 sessions/week). FAST training consisted of walking-related activities for 40 minutes/session at 70-80% heart rate reserve, while SAM received daily feedback and goal-setting of walking activity (steps per day). Assessors and study statistician were masked to group assignment.

The *a priori*-determined primary outcome and primary endpoint was change in steps per day from pre- to post-intervention. Adverse events (AEs) were tracked after randomization. All randomized participants were included in the intent-to-treat analysis. This study is registered at ClinicalTrials.gov, NCT02835313.

**Findings:** Participants were enrolled from July 18, 2016-November 16, 2021. Of 250 randomized participants (mean[SE] age 63[0.80], 116F/134M), 89 were assigned to FAST, 81 to SAM, and 80 to FAST+SAM. Steps per day significantly increased in both the SAM (mean[SE] 1542[267], 95%CI:1014-2069, *p*<0.001) and FAST+SAM groups (1307[280], 752-1861, *p*<0.001), but not in the FAST group (406[238], 63-876, *p*=0.09).

There were no deaths or serious study-related AEs and all other minor AEs were similar between groups.

**Interpretation:** Only individuals with chronic stroke who completed a step activity monitoring behavioral intervention with skilled coaching and goal progression demonstrated improvements in physical activity (steps per day).

## INTRODUCTION

Despite advances in stroke interventions and rehabilitation, the worldwide burden of post-stroke disability is growing. ^1–3^ The total global costs of stroke are ∼ US$891 billion^4^ with a 143% increase in disability-adjusted life-years lost. ^5^ Six to twelve months after stroke, over 50% of survivors have persistent deficits leading to impaired walking and decreased independence. ^6^ This stroke-related disability is both a consequence of, and risk factor for, physical inactivity, ^6–8^ and places individuals at an increased risk of cardiovascular disease, diabetes, additional stroke(s), myocardial infarction, or death. ^1, 6, 7, 9–12^ Individuals after stroke who are inactive have more than three times the risk of recurrent stroke as their more active peers. ^1^ Thus, there is a complex interplay between lack of physical activity, disability and poor health post-stroke, and improving physical activity is an important target for breaking this vicious cycle.

Daily walking activity, as measured by steps per day, is a highly relevant method of quantifying physical activity for individuals with chronic stroke, as increased daily walking has been associated with healthy values of blood pressure and body weight and lower rates of cardiovascular disease and diabetes. ^13–15^ Increasing walking activity, therefore, has the potential to directly counter some of the devastating health complications that are prevalent post-stroke.

In the World Health Organization’s International Classification of Functioning, Disability and Health (ICF) model, there are 2 qualifiers in the Activity domain – capacity and performance. ^16^ Capacity describes what someone is capable of doing as assessed in a standardized environment such as a clinic or laboratory. Performance describes the activity that a person actually does in their unstructured, everyday environment. Despite evidence that specific rehabilitation interventions can improve walking <u>capacity</u> post-stroke (e.g., distance covered on the six-minute walk test, walking speed), ^17, 18^ concomitant improvements in walking <u>performance</u> (e.g. walking activity as measured by steps per day in the free-living environment), are generally not observed ^19–21^ or are relatively modest. ^22^ This lack of translation of gains in walking capacity to gains in walking performance is well-documented post-stroke and is also observed in upper extremity tasks. ^23^

Given the lack of translation of gains in walking capacity to gains in walking performance, rehabilitation interventions may need to include strategies that aim to improve both walking capacity (distance, speed), <u>and</u> walking performance (steps per day). For improving walking capacity in the chronic stages post-stroke (>6 months duration), recent work has highlighted the comparative efficacy of high-intensity walking training. ^17, 18^ For improving walking performance, results from studies in other health conditions and one small study in individuals with chronic stroke demonstrate that behavioral interventions with step activity monitoring, which include tracking walking activity with an accelerometer and goal setting, may be effective in increasing steps per day. ^24, 25^ These collective results suggest that combining a high-intensity walking training program designed to improve walking capacity with a step monitoring program designed to improve walking performance may result in improvements in both walking capacity and walking performance post-stroke. To test this theory, we previously completed a feasibility and preliminary efficacy study combining these interventions in people with chronic stroke. ^26^ The results of this preliminary study showed that those with lower baseline steps per day had greater improvements in steps per day with the combined intervention compared to the high-intensity walking intervention alone. ^26^ In the present assessor blinded, multi-site, randomized controlled trial, we aimed to extend the previous evidence by examining the individual and combined effects of a high-intensity walking intervention and step activity monitoring behavioral intervention on steps per day in people with chronic stroke. We hypothesized the combined program would yield a greater increase in steps per day than either intervention alone.

## METHODS

The Promoting Recovery Optimization with WALKing Exercise after Stroke (PROWALKS) study was a multi-site assessor-blind randomized controlled trial conducted in university/hospital-based physical therapy laboratories at the University of Delaware, Christiana Care Health System, University of Pennsylvania, and Indiana University. Participants were enrolled from July 2016-November 2021. The *a priori* determined primary outcome and endpoint was change in steps per day assessed prior to randomization and following the completion of the intervention. The study was approved by all sites’ institutional review boards, and the protocol has been published. ^8^

### Participants

Participants were recruited from the community through clinicians and support groups, advertisement, existing databases, and health record systems. Participants were enrolled prior to the initial PRE evaluation (before randomization) by the assessor physical therapist at each site. Participants provided written informed consent and met the inclusion criteria as follows: 1) age 21-85 years, 2) ≥6 months post-stroke, 3) able to walk without assistance of another person (assistive devices and orthotics permitted), 4) self-selected walking speeds 0.3 m/s-1.0 m/s, 5) average steps per day <8,000, 6) resting heart rate (HR) 40-100 beats per minute, 7) resting blood pressure (BP) 90/60-170/90. Exclusion criteria included: 1) evidence of cerebellar stroke, 2) other neurological deficits unrelated to stroke, 3) lower limb botulinum toxin injection ≤4 months previous, 4) currently in physical therapy, 5) unable to walk outside the home prior to stroke, 6) coronary artery bypass graft, stent placement, or myocardial infarction ≤3 months previously, 7) inability to communicate with investigators, 8) score >1 on question 1b and >0 on question 1c on the NIH Stroke Scale (indicating the individual was not oriented and/or could not follow simple commands) (See Figure 1). ^8^ The gait speed lower limit of 0.3 m/s was established shortly after the start of the study, due to challenges with accuracy of Fitbit devices when gait speed was slower than 0.3 m/s. ^27^ Participants provided self-reported sex, race and ethnicity data and this information was included to describe the study sample.

**Figure 1.**
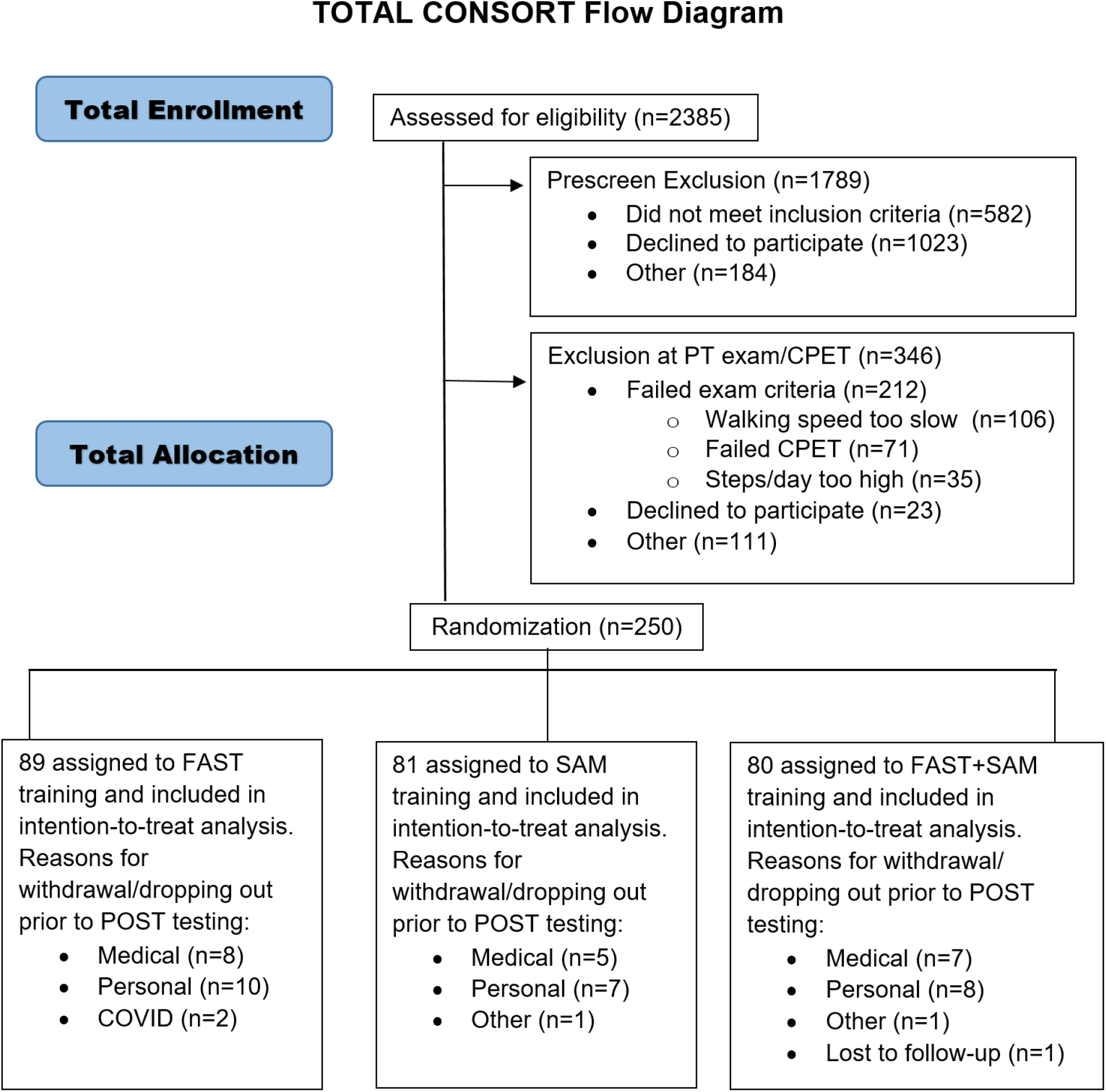
CONSORT diagram. PT denotes physical therapy.

### Outcome Assessment

Outcomes were assessed at each site by blinded evaluators before randomization (PRE) and following the end of the intervention (POST). The outcome assessment procedures were the same for participants in all groups. The primary outcome quantified walking performance via steps per day, assessed by the Fitbit^TM^ One or Fitbit^TM^ Zip. The step monitor was worn on the non-paretic ankle for 7 full days, with a minimum of 3 valid recordings days included in analysis. ^27–30^ Data was retrieved for analysis through the Fitabase^TM^ platform. Secondary outcomes of walking capacity included distance walked on the 6-minute walk test (6MWT), ^31^ self-selected walking speed during the 10-meter walk test (SSWS), ^31^ and oxygen consumption (VO_2_ at ventilatory threshold) as measured on a maximal treadmill cardiopulmonary exercise test (CPET). ^8, 15^

### Randomization and Masking

Following screening and PRE testing, 250 participants were randomly assigned to one of the following groups: ^8^ 1) high-intensity walking alone (FAST), 2) step activity monitoring behavioral intervention alone (SAM), or 3) combined high-intensity walking and step activity monitoring behavioral intervention (FAST+SAM) (Figure 1). Participants were assigned using a random allocation sequence and concealed allocation. All allocation sequences were generated by the study coordinator using https://www.randomizer.org. Group assignment was kept in sequentially numbered sealed opaque envelopes opened by the study coordinator and communicated directly to the training physical therapist. Separate random allocation sequences were created for each study site. The study coordinator who made and communicated group assignments did not have any involvement in training, assessments, or data analysis. The biostatistician (RP) was blinded until primary and secondary outcome analyses were completed.

### Adverse Event Monitoring

At each study visit, participants were asked if they experienced adverse events (AEs) including pain, falls, or illness, since the previous session. During testing and training, participants’ vital signs and other signs or symptoms of cardiorespiratory insufficiency, orthopedic injury or other issues were monitored, using accepted stopping criteria as defined at the study start including American College of Sports Medicine recommendations. ^8^ AEs were categorized as minor/serious, expected/unexpected, and study-related/not related as designated prior to the start of the study. ^8^ A data safety monitoring board was in place for the duration of the study.

### Training Interventions

The attendance goal for all 3 groups was up to 36 sessions (∼3x/week for 12 weeks). Participants receiving FAST training completed up to 30 minutes/session of treadmill walking followed by 10 minutes of overground activities, all while maximizing time within their Target Heart Rate (THR) range (70-80% heart rate reserve). ^8^ SAM training included individualized goal-setting for steps per day and coaching to improve daily walking activity.

#### FAST walking intervention (FAST and FAST+SAM groups)

All treadmill walking was completed with an overhead chest-harness system for safety, which provided no body weight support. Participants were allowed to use handrails as needed throughout treadmill walking. Treadmill incline was adjusted, or weight vests or ankle weights added as needed to keep heart rate in the THR range during all walking. The treadmill speed was reduced if the: 1) participant requested to walk slower, 2) speed was no longer safe (e.g. increased toe scuffing or tripping), 3) participant reported a rate of perceived exertion (RPE) of ≥17 on the Borg RPE Scale, ^32^ or 4) THR range was exceeded. If HR was exceeded, treadmill speed was lowered until HR returned to 50-60% heart rate reserve ^33^ or RPE ≤13, at which time treadmill speed was increased to bring the participant back into the THR range.

Overground activities were performed with the participant’s usual assistive device. The goal was to practice important everyday community walking activities while maximizing time spent in the THR range (e.g. turning, backward stepping, stair climbing, carrying objects). Participants were guarded by the physical therapist during all overground walking, and these activities progressed throughout training.

#### Step activity monitoring behavioral intervention (SAM and FAST+SAM groups)

Participants were provided with a Fitbit to track steps per day. Therapists used step activity data from PRE to determine the initial individualized step activity goal in consultation with the participant. Participants were encouraged to achieve their step goal each day. Therapists used the Fitbit application to review a participant’s step activity between sessions. At each in-person session, participants and therapists discussed whether their average steps per day since the prior session reached their step activity goal and on which days this occurred. If the step activity goal was achieved regularly, the participant and therapist together determined an appropriate increase in the goal. The objective was an increase in the step activity goal by 5-8% every 4-6 sessions. If the step activity goal was not met, individualized strategies were identified to increase walking activity and overcome barriers using a motivational interviewing framework. ^34, 35^

### Statistical Analyses

Study sample size, power, and statistical plan were previously published in our protocol paper. ^8^ Briefly, an n of 225 (75 per group) would allow us to detect a difference of 1700 steps per day over time among intervention groups, with power ≥ 0.90, and α=0.05. ^8^ To account for attrition, 250 participants were randomized into the three groups. The study was prospectively registered at ClinicalTrials.gov (NCT02835313). Consistent with RCT standards, our analysis used an intent-to-treat approach. Initial analyses were conducted blind to group membership and data were analyzed using SPSS version 27 (IBM, Chicago, IL, USA).

To ensure groups were balanced participant characteristics were compared across groups at baseline using one-way ANOVAs for continuous variables and Fisher exact tests for categorical variables (Table 1).

**Table 1.** Baseline participant characteristics.

| **BASELINE PARTICIPANT CHARACTERISTICS (either by mean (SE) or N (%))** |  |  |  |
| --- | --- | --- | --- |
|  | **FAST alone**<br>**(n=89)** | **SAM alone**<br>**(n=81)** | **FAST + SAM**<br>**(n=80)** |
| Age, years | 63.98 (1.27) | 62.16 (1.44) | 62.10 (1.46) |
| Time since stroke, months | 46.12 (6.60) | 46.77 (8.14) | 42.73 (5.45) |
| Body mass index | 29.79 (0.69) | 31.58 (0.78) | 30.13 (0.61) |
| Self-selected gait speed, m/s | 0.71 (0.02) | 0.68 (0.02) | 0.68 (0.03) |
| 6-minute walk test distance, m | 296 (12) | 279 (12) | 283 (14) |
| Steps per day | 3959 (213) | 3845 (242) | 3768 (232) |
| VO<sub>2</sub> at ventilatory threshold (mL/kg/min) | 8.77 (0.31) | 9.37 (0.26) | 9.63 (0.30) |
| Females, N (%) | 41 (46.07%) | 39 (48.15%) | 36 (45.00%) |
| Race, N (%) |  |  |  |
| Native American/Alaskan native | 4 (4.49%) | 4 (4.94%) | 3 (3.75%) |
| Asian | 8 (8.99%) | 2 (2.47%) | 4 (5.00%) |
| Native Hawaiian/Pacific islander | 0 | 0 | 0 |
| Black/African-American | 24 (26.97%) | 27 (33.33%) | 21 (26.25%) |
| White | 57 (64.04%) | 52 (64.20%) | 55 (68.75%) |
| Multiple races | 5 (5.62%) | 5 (6.17%) | 6 (7.50%) |
| Prefer not to identify race | 1 (1.12%) | 0 | 1 (1.25%) |
| Hispanic/Latinx ethnicity, N (%) | 5 (5.62%) | 2 (2.57%) | 3 (3.75%) |
| Side of hemiparesis, N (%) |  |  |  |
| Left | 43 (48.31%) | 42 (51.85%) | 39 (48.75%) |
| Right | 43 (48.31%) | 34 (41.98%) | 37 (46.25%) |
| Both affected | 3 (3.37%) | 5 (6.17%) | 4 (5.00%) |
| Assistive device use, N (%) | 46 (51.69%) | 47 (58.02%) | 36 (45.00%) |
| Orthotic device use, N (%) | 21 (23.60%) | 25 (30.86%) | 22 (27.50%) |

To ensure that interventions were equivalently delivered, a one-way ANOVA was used to compare the number of training sessions completed for each group. Additionally, the average number of minutes walking on the treadmill per session and percentage of treadmill time in the THR range were compared between the FAST and FAST+SAM groups. The average number of Fitbit-wearing days per week and percentage of Fitbit-wearing days on which the step goal was achieved were compared between the SAM and FAST+SAM groups.

Marginal General Linear Mixed Models were used for the primary and secondary outcomes. Fixed effects included the main effects of group and time, along with their interactions; a compound symmetric covariance matrix for the errors was used to account for the repeated measures. This was chosen by finding the best fitting covariance matrix as determined by examining the Akaike information criterion (AIC) and Bayesian information criterion (BIC). The Satterthwaite approximation was used for determining degrees of freedom. The models used robust errors (Hubert-White) to account for mild departures of normality. Maximum likelihood estimation was employed to garner estimates in the presence of missing data without needing listwise deletion. Study site was attempted to be included as a random effect, but this resulted in model non-convergence, likely due to the lower number of sites (4); therefore, additional models were run testing site as a fixed effect. As it was non-significant for all outcomes, the results are reported without it.

## RESULTS

A total of 2385 individuals were assessed for eligibility (Figure 1); 2135 were excluded prior to randomization, and the remaining 250 were randomized. The baseline characteristics of the participants were similar (Table 1). The participants had a mean [SE] age of 62 [0.80], years and were an average of 45 [3.9] months post-stroke. The race, ethnicity, hemiplegic side, assistive device and orthotic use, and clinical performance were balanced between groups. Since none of the potentially clinically-relevant covariates were found to be different among groups at baseline (*p*=0.16-1.00 for potential covariates), none were included in the analysis model (Table 1).

Between-group comparisons of training variables were performed to test whether there were differences in training to assess treatment fidelity. These variables are presented in Table 3. There were no between-group differences for number of sessions attended (*p*=0.45), average treadmill walking minutes/session (*p*=0.68), percentage of treadmill minutes in THR range (*p*=0.62), or percentage of Fitbit-wearing days on which goals were achieved (*p*=0.31). The FAST+SAM group wore their Fitbits significantly fewer days/week than the SAM group (3.96 versus 4.40 days/week, *p*=0.002; 95% CI=0.18-0.83).

For the primary outcome (steps per day), the change over time between groups was different (*p*=0.004) (Table 2, Figure 2). The SAM and FAST+SAM group significantly increased their steps per day from PRE to POST, including an increase of 1542 steps (95% CI=1014-2069, *p*<0.001) for SAM, and an increase of 1307 steps (95% CI=752-1861, *p*<0.001) for FAST+SAM. Conversely, the FAST group only increased steps per day by 406 (95% CI=-63-876, *p*=0.09). The difference in steps per day change between the SAM and FAST+SAM groups was not significant (*p*=0.58).

**Figure 2.**
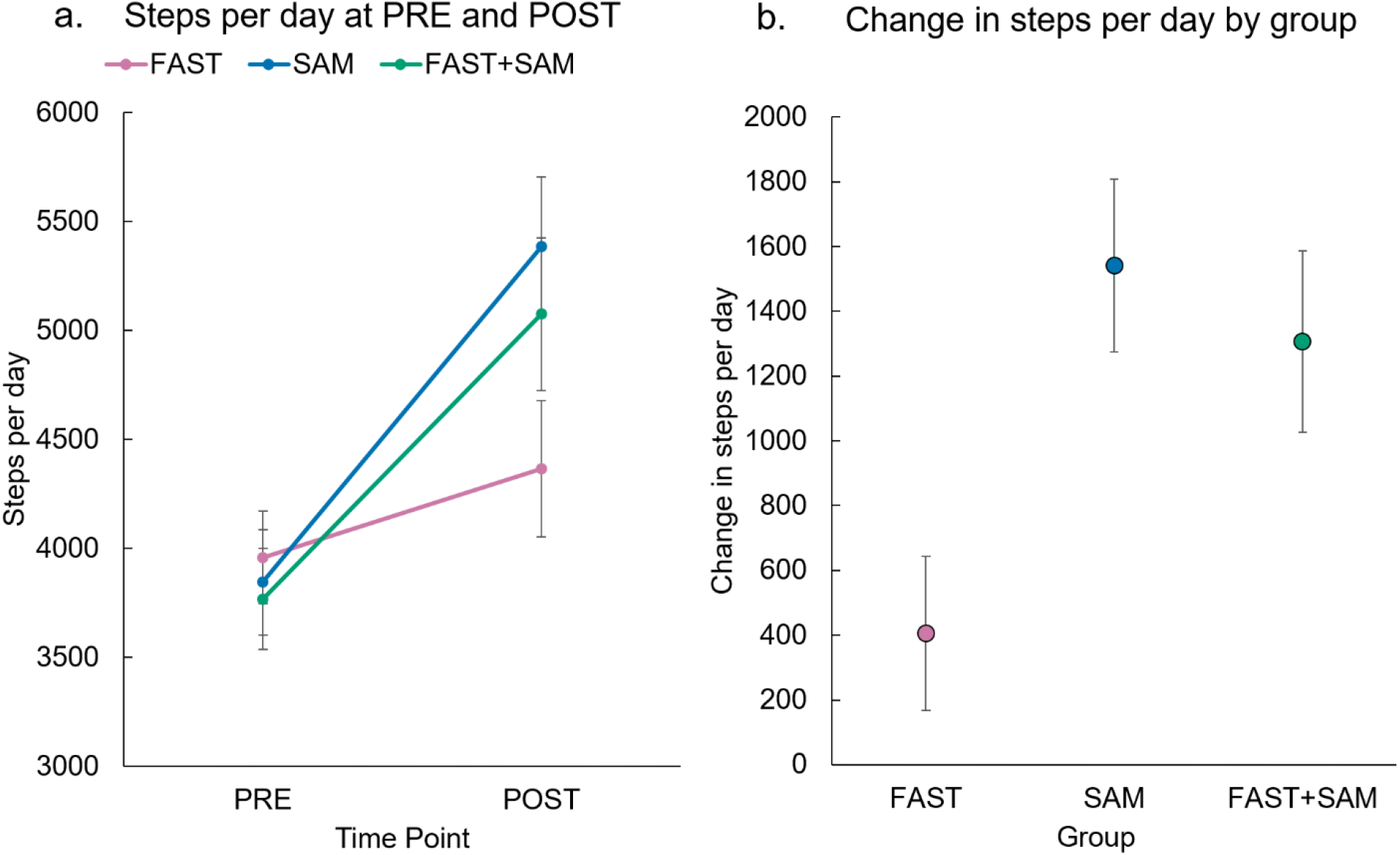
Primary outcome: Change in steps per day by group. a) Steps per day mean values by group (error bars denote standard error). SAM and FAST+SAM groups increased significantly (*p*<0.001 for each group), while FAST alone did not (*p*=0.09). b) Magnitude of steps per day change for each group (as mean(SE)).

**Table 2.** Change in study outcomes (primary = steps per day; secondary = 6MWT distance, self-selected walking speed, VO_2_ at ventilatory threshold) by group, including mean, standard error, 95% confidence interval for change between PRE and POST. (Abbreviations: SE = standard error; CI = confidence interval; 6MWT = 6-minute walk test; SSWS = self-selected walking speed; VO_2_ = oxygen consumption)

| **OUTCOME CHANGES BETWEEN PRE AND POST (mean (SE) [95% CI for change])** |  |  |  |
| --- | --- | --- | --- |
| **Outcome** | **FAST alone** | **SAM alone** | **FAST + SAM** |
| Steps per day |  |  |  |
| PRE | 3959 (213) | 3845 (242) | 3768 (232) |
| POST | 4365 (311) | 5386 (319) | 5075 (351) |
| Change (POST-PRE) | 406 (238) | 1542 (267) | 1307 (281) |
|  | [-63-876] | [1014-2069] | [752-1861] |
| 6MWT distance (m) |  |  |  |
| PRE | 296 (12) | 279 (12) | 283 (14) |
| POST | 338 (13) | 304 (13) | 323 (15) |
| Change (POST-PRE) | 43 (6) | 25 (6) | 39 (8) |
|  | [32-54] | [13-36] | [23-56] |
| SSWS (m/s) |  |  |  |
| PRE | 0.71 (0.02) | 0.68 (0.02) | 0.68 (0.03) |
| POST | 0.84 (0.03) | 0.79 (0.03) | 0.82 (0.03) |
| Change (POST-PRE) | 0.13 (0.02) | 0.11 (0.02) | 0.14 (0.02) |
|  | [0.10-0.17] | [0.08-0.14] | [0.10-0.17] |
| VO<sub>2</sub> (mL/kg/min) |  |  |  |
| PRE | 8.77 (0.31) | 9.37 (0.26) | 9.63 (0.30) |
| POST | 9.04 (0.34) | 9.38 (0.34) | 9.32 (0.38) |
| Change (POST-PRE) | 0.27 (0.39) | 0.01 (0.37) | 0.31 (0.42) |
|  | [-1.05-0.50] | [-0.73-0.71] | [-0.52-1.13] |

**Table 3.**
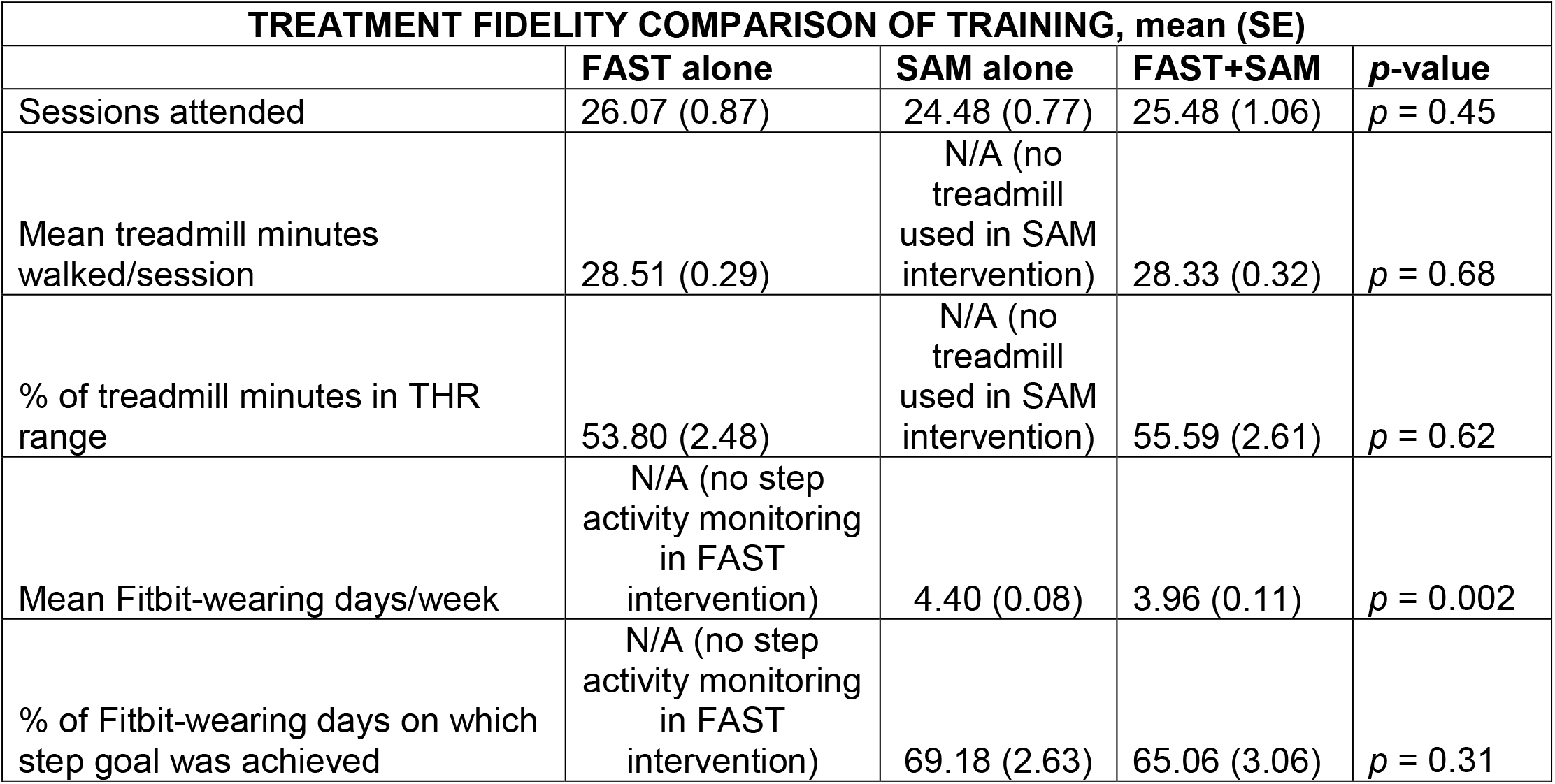
Treatment fidelity comparison of training in single-intervention groups (FAST, SAM) versus the combined group (FAST+SAM). Only one between-group difference was found: participants in the FAST+SAM group wore their Fitbits significantly fewer days per week during training than those in the SAM group. (Abbreviations: SE = standard error; N/A = not applicable)

| **TREATMENT FIDELITY COMPARISON OF TRAINING, mean (SE)** |  |  |  |  |
| --- | --- | --- | --- | --- |
|  | **FAST alone** | **SAM alone** | **FAST+SAM** | **p-value** |
| Sessions attended | 26.07 (0.87) | 24.48 (0.77) | 25.48 (1.06) | $p = 0.45$ |
| Mean treadmill minutes walked/session | 28.51 (0.29) | N/A (no treadmill used in SAM intervention) | 28.33 (0.32) | $p = 0.68$ |
| % of treadmill minutes in THR range | 53.80 (2.48) | N/A (no treadmill used in SAM intervention) | 55.59 (2.61) | $p = 0.62$ |
| Mean Fitbit-wearing days/week | N/A (no step activity monitoring in FAST intervention) | 4.40 (0.08) | 3.96 (0.11) | $p = 0.002$ |
| % of Fitbit-wearing days on which step goal was achieved | N/A (no step activity monitoring in FAST intervention) | 69.18 (2.63) | 65.06 (3.06) | $p = 0.31$ |

For 6MWT distance, all groups improved from PRE to POST (*p*<0.001 for each group, including a 43m, (95% CI=32-54) increase for FAST; a 25m (95% CI=13-36) increase for SAM, and a 39m, (95% CI=23-56) increase for FAST+SAM. All groups also improved their speeds during the 10 MWT from PRE to POST (*p*<0.001 for each group), including a 0.13m/s (95% CI=0.10-0.17) increase for FAST; a 0.11m/s, (95% CI=0.08-0.14) increase for SAM, and an increase of 0.14m/s, (95% CI=0.10-0.17) for FAST+SAM. For VO_2_, there were no effects of time or group (*p*=0.96, *p*=0.12, respectively).

There were no serious study-related AEs between PRE and POST (Table 4). Seven minor study-related AEs were reported (muscle pain/soreness (n=5), dizziness/fainting (n=1) and falls (n=1)). Eleven serious non-study-related AEs were reported (fracture (n=1) and hospitalizations (n=10)). The majority of AEs were minor and non-study-related (n=63), (falls (n=43), muscle pain/soreness (n=10), COVID-19 (n=3) or other illness (n=7)). Non-study-related totals for falls and muscle pain/soreness reflect some participants sustaining the same event multiple times.

**Table 4.**
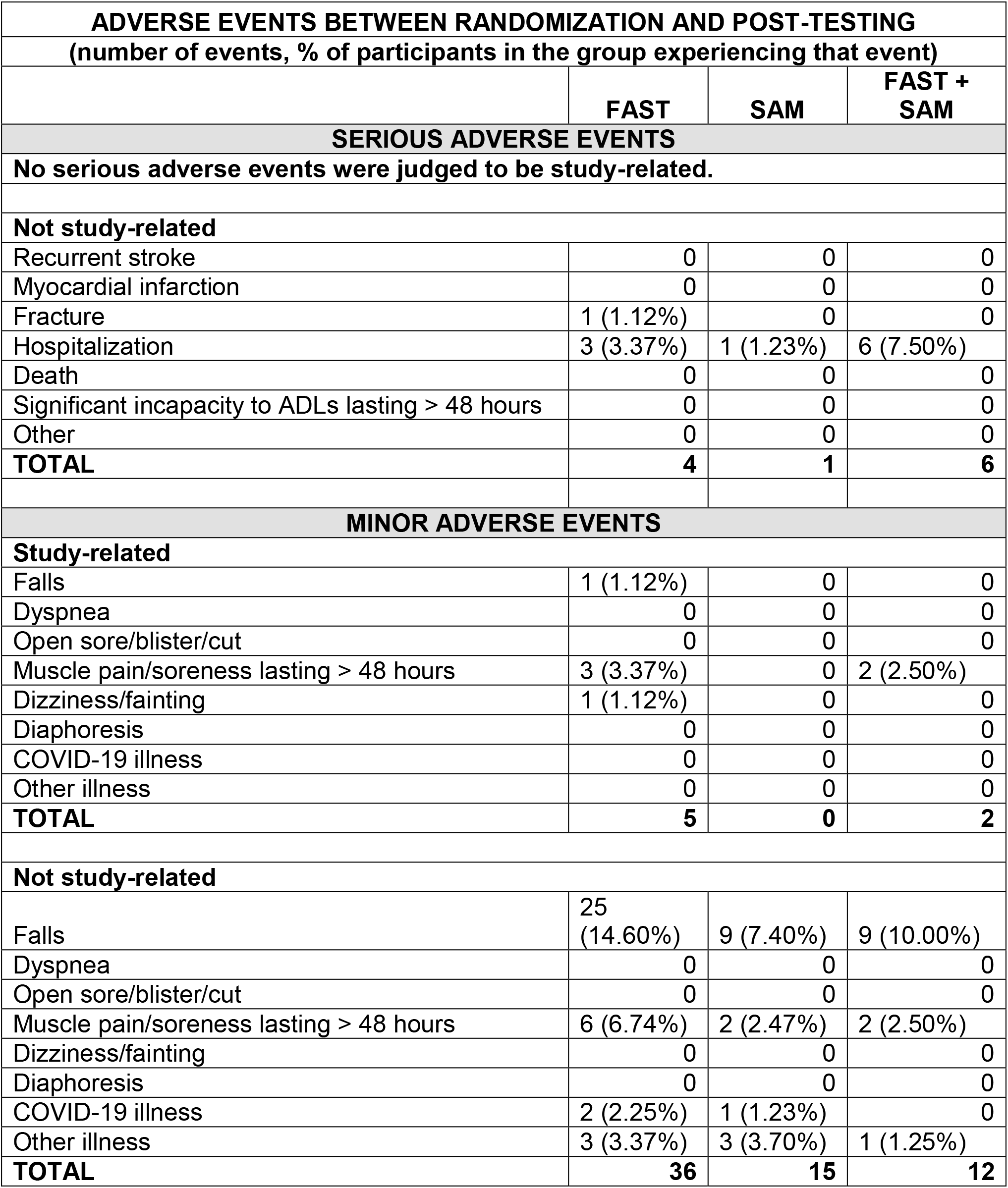
Adverse events by group. Due to some participants experiencing falls or muscle pain/soreness multiple times, all data are presented both as the raw number of events per category, and the percentages of participants in each group who experienced that event. With non-study-related falls, for example, 14 falls in the FAST group were sustained by five people, seven falls in the SAM group were sustained by three people, and two falls in the FAST+SAM group were sustained by one person. (Abbreviations: ADLs = activities of daily living)

| **ADVERSE EVENTS BETWEEN RANDOMIZATION AND POST-TESTING** |  |  |  |
| --- | --- | --- | --- |
| **(number of events, % of participants in the group experiencing that event)** |  |  |  |
|  | **FAST** | **SAM** | **FAST + SAM** |
| **SERIOUS ADVERSE EVENTS** |  |  |  |
| **No serious adverse events were judged to be study-related.** |  |  |  |
| **Not study-related** |  |  |  |
| Recurrent stroke | 0 | 0 | 0 |
| Myocardial infarction | 0 | 0 | 0 |
| Fracture | 1 (1.12%) | 0 | 0 |
| Hospitalization | 3 (3.37%) | 1 (1.23%) | 6 (7.50%) |
| Death | 0 | 0 | 0 |
| Significant incapacity to ADLs lasting > 48 hours | 0 | 0 | 0 |
| Other | 0 | 0 | 0 |
| **TOTAL** | **4** | **1** | **6** |
| **MINOR ADVERSE EVENTS** |  |  |  |
| **Study-related** |  |  |  |
| Falls | 1 (1.12%) | 0 | 0 |
| Dyspnea | 0 | 0 | 0 |
| Open sore/blister/cut | 0 | 0 | 0 |
| Muscle pain/soreness lasting > 48 hours | 3 (3.37%) | 0 | 2 (2.50%) |
| Dizziness/fainting | 1 (1.12%) | 0 | 0 |
| Diaphoresis | 0 | 0 | 0 |
| COVID-19 illness | 0 | 0 | 0 |
| Other illness | 0 | 0 | 0 |
| **TOTAL** | **5** | **0** | **2** |
| **Not study-related** |  |  |  |
| Falls | 25<br>(14.60%) | 9 (7.40%) | 9 (10.00%) |
| Dyspnea | 0 | 0 | 0 |
| Open sore/blister/cut | 0 | 0 | 0 |
| Muscle pain/soreness lasting > 48 hours | 6 (6.74%) | 2 (2.47%) | 2 (2.50%) |
| Dizziness/fainting | 0 | 0 | 0 |
| Diaphoresis | 0 | 0 | 0 |
| COVID-19 illness | 2 (2.25%) | 1 (1.23%) | 0 |
| Other illness | 3 (3.37%) | 3 (3.70%) | 1 (1.25%) |
| **TOTAL** | **36** | **15** | **12** |

## DISCUSSION

The results of the PROWALKS study showed that walking activity, as measured by steps per day, increased significantly in the SAM and FAST+SAM groups, but not the FAST group. The data demonstrate that gains in walking activity in those with chronic stroke occur following a targeted behavioral intervention integrating a step monitoring program with skilled coaching and goal progression, and not following a high intensity walking intervention alone. These results provide an important step forward in our understanding of the interventions necessary to improve walking activity after stroke by demonstrating that increasing walking capacity alone is not sufficient for improving walking activity in the real world after stroke.

Physical inactivity after stroke can have devastating consequences related to disability and health, including decreased functional independence and increased risk of recurrent stroke and mortality. ^1, 6, 7, 9–12^ Recent systematic reviews and meta-analyses of physical activity in people with stroke suggest profound inactivity in this population. ^36, 37^ Across 1280 participants across 32 studies, it was found that individuals with chronic stroke only perform 4078 steps per day. While there is no published minimal clinically important difference (MCID) for steps per day in people with chronic stroke, the average improvements in steps per day found in the SAM (1542) and FAST+SAM (1307) groups in this study represent an increase of more than 30% of the steps normally taken by people with chronic stroke. ^36, 37^ These changes in steps per day are almost double the typical changes (0-650 steps per day) that have been found with previous walking interventions in people with chronic stroke. ^19, 20, 22, 38^ Given the complex interplay between lack of physical activity, disability and poor health status after stroke, the substantial gains in walking activity found in those in the SAM and FAST+SAM group have important implications for people living with stroke.

In contrast to the results for the SAM and FAST+SAM groups, the change in steps per day in the FAST group (406) is consistent with prior studies of moderate to high-intensity walking interventions after stroke that did not incorporate a step activity monitoring behavioral intervention. These studies found either no change ^19–21^ or relatively small gains that are comparable to the FAST group in this study. ^22, 38, 39^ Mirroring the results of the FAST group, a previous study found that gains in walking speed following outpatient rehabilitation after stroke were not accompanied by gains in steps per day. ^23^

Relating these findings back to the domains of capacity and performance, the present and previous results reinforce the idea that targeting and improving walking capacity alone may not be sufficient to observe changes in walking performance post-stroke. This lack of translation of improvements in capacity to improvements in performance is also observed with upper extremity interventions in people with stroke. ^23^ Thus, it appears that this disconnect could be a general feature of post-stroke recovery. If so, a potential paradigm shift in post-stroke rehabilitation may be warranted. Post-stroke rehabilitation interventions may need to be specifically designed to target improvements in performance in the real world, rather than relying primarily on interventions that target improvements in capacity to directly translate to performance.

When comparing groups on secondary outcomes, participants in the FAST+SAM and FAST groups saw similar improvements in 6MWT distance, (39m and 43 respectively), with smaller changes observed in participants in the SAM group (25m). The gains in FAST+SAM and FAST exceed the MCID range for a moderate change in 6MWT in those with chronic stroke or related populations (34-50m^40–42^) and the minimal detectable change (MDC_90_) for 6MWT in those with chronic stroke (34-43m^31, 43^), whereas the SAM group’s change falls below these thresholds. Notably, 61% and 49% of participants in FAST+SAM and FAST, respectively, surpassed the threshold MCID whereas, following SAM training, only 28% of participants surpassed this threshold. The lower changes in 6MWT distance in the SAM group may be the result of not participating in an intervention specifically aimed at improving walking capacity. This further reinforces the idea that interventions post-stroke may need to be targeted specifically at improving walking capacity and/or walking performance.

The results for walking speed and oxygen consumption demonstrated no differences between groups. Walking speed significantly improved across all groups, and exceeded the commonly used MCID threshold of 0.1 m/s, ^44^ while oxygen consumption did not change. This lack of change in VO_2_ could be due to the changes in gait speed, as improvements in gait speed can result in improved gait efficiency post-stroke. ^45^ This has confounding effects on VO_2_ during exercise testing because improved gait efficiency may result in a decrease in measured VO_2_. ^46^

Only participants in the FAST + SAM group experienced a clinically relevant improvement in steps per day, 6MWT distance, and walking speed. This combined change in walking performance and walking capacity was not demonstrated by either single-intervention group and suggests that a combined intervention may be best for addressing deficits in <u>both</u> walking capacity and performance in those with chronic stroke. However, our hypothesis that the FAST+SAM group would improve steps per day more than either the FAST or SAM intervention alone was only partially supported by these results. FAST+SAM did not show a greater increase in steps per day than SAM, despite the additional walking capacity-building training that the participants in FAST+SAM received. Based on previous findings, ^26^ it is possible that only certain sub-groups of people with chronic stroke, such as those with low baseline walking capacity or steps per day, will show preferential benefit from an intervention that targets both capacity and performance.

### Limitations

Limitations of the study include a sample size which was calculated to detect a significant change in steps per day at 90% power; this sample size may have limited our ability to detect changes among our secondary outcomes. How well the results might generalize to people with chronic stroke who walk slower than 0.3 m/s or who require physical assistance from another person to walk is unclear. Finally, given the heterogeneity in this population, further planned analyses will identify characteristics of participants who benefited more from each intervention in terms of walking performance, walking capacity, or both.

### Conclusions

Individuals with chronic stroke who completed a program emphasizing increased daily activity through step monitoring, individualized goals, and coaching demonstrated significant improvements in physical activity, as measured by steps per day. Only the group that received both the high intensity walking intervention and the step activity monitoring behavioral intervention achieved a clinically meaningful improvement in walking performance (steps per day) <u>and</u> in both measures of walking capacity (6MWT distance, walking speed). These findings demonstrate that a step activity monitoring behavioral intervention results in large changes in walking activity in people with chronic stroke, which could decrease the risk of costly and debilitating complications, including cardiac events, loss of independence, and mortality.

## ACKNOWLEDGMENTS AND SOURCE OF FUNDING

This study was funded by the National Institutes of Health, grant # NIH-R01HD086362.

## Data Availability

All data produced in the present study are available upon reasonable request to the authors

